# Reducing friction, enabling response: a realist evaluation of a mobile outreach model for marginalized populations

**DOI:** 10.64898/2026.07.30.26358580

**Authors:** Sayra Cristancho, Don Eby, Francesca Dobbyn, Kevin McNab

## Abstract

**Background:** Mobile outreach initiatives have emerged to address persistent barriers to care for people experiencing homelessness, substance use, and mental illness. Although these models show promise, less is known about how and under what conditions they enable engagement and coordinated care. This study explains how, why, and under what circumstances a mobile, cross-sector outreach model enables access to care for marginalized populations.

**Methods:** We conducted a realist evaluation of Supportive Outreach Services (S.O.S.), a mobile, cross-sector outreach program in Grey County, Ontario. Data included 31 semi-structured interviews with outreach providers, partner organizations, system leaders, and clients, supplemented by document review and stakeholder feedback. Using retroductive reasoning and constant comparison, we developed and refined context–mechanism–outcome configurations to construct an explanatory program theory.

**Results:** Five interconnected realist explanations account for how the model enables access to care. Trust built through repeated, non-judgmental encounters supports engagement; proximity reduces barriers to participation; accessible support enables timely help-seeking; cross-sector relationships enable adaptive coordination; and visible results build legitimacy that sustains participation and resources. Together, these explanations provide a linked explanatory account of how mobile outreach reduces friction between marginalized populations and fragmented services while identifying the structural conditions that constrain its effectiveness.

**Conclusions:** The effectiveness of mobile outreach depends less on the services delivered than on its capacity to reduce friction, sustain relationships, and adapt care across organizational boundaries. The resulting program theory offers transferable explanations for designing coordinated community-based services while highlighting the structural conditions required for durable change.

## INTRODUCTION

Health and social care systems continue to struggle to engage individuals experiencing homelessness, substance use, and mental illness—particularly in smaller communities where services are limited and fragmented.^1-2^ Barriers are well documented: stigma, rigid eligibility criteria, geographic distance, and the cumulative burden of navigating disconnected systems.^3-4^ These conditions often result in delayed care, reliance on emergency services, and poor health outcomes.^5-6^

In response, community-initiated models have emerged that seek to bring care to people rather than requiring people to access care through conventional pathways. Supportive Outreach Services (S.O.S.) is one such model.^7^ Developed in Grey County, Ontario, S.O.S. is a mobile, cross-sector initiative that integrates health care, harm reduction, and social support, delivered directly in the environments where people live. It operates through a combination of mobility, relationship-based engagement, and collaboration across organizations.

Rather than replicating existing services, S.O.S. reconfigures how care is accessed and coordinated. It prioritizes low-barrier entry, sustained presence, and flexible, cross-sector responses to complex needs. Early descriptive work has highlighted its emphasis on trust-building, harm reduction, and partnerships as central to its functioning.^8^ What remains less understood is how these elements work together to produce outcomes, and under what conditions they are effective.

A realist evaluation is well suited to this question.^9-10^ Realist approaches move beyond asking whether an intervention works to examine how, why, and under what circumstances outcomes are generated, through the interaction of context-mechanism-outcome configurations (CMOCs).^11^ This study applies a realist lens to develop an explanatory account of how S.O.S. enables engagement, reduces harm, and supports access to coordinated care.^12^

The aim of this study is to explain how, why, and under what conditions a mobile, cross-sector outreach model enables access to care for marginalized populations. By identifying the mechanisms through which S.O.S. operates – and the contexts that sustain or constrain them – this study seeks to inform the design of coordinated service delivery models in similarly resource-constrained settings.

## METHODS

### Design

We conducted a realist evaluation to develop an explanatory account of how S.O.S. produces outcomes in practice. Realist evaluation is a theory-driven approach that seeks to explain what works, for whom, in what circumstances, and why, by identifying context– mechanism–outcome configurations (CMOCs).^9-11^

### Study setting

S.O.S. is a mobile, cross-sector initiative operating in Grey County, Ontario. It brings together providers from health care, social services, paramedicine, and charitable organizations to deliver care directly in community settings, including streets, encampments, shelters, and rural locations.^8^

### Data collection

Data were drawn from a qualitative study conducted between January and June 2025. The study included 31 semi-structured interviews with a purposive sample of participants, including members of the mobile team, partner organizations, system leaders, and clients. Interviews explored experiences of delivering and receiving care within S.O.S., with attention to how the model functioned in practice.

Interviews were audio-recorded, transcribed verbatim, and supplemented with document review and stakeholder feedback sessions used to refine emerging interpretations.

### Analysis

Analysis followed a realist logic of inquiry. Data were coded iteratively to identify recurring patterns linking contexts, mechanisms, and outcomes. Through a process of constant comparison and retroductive reasoning, we developed and refined 47 CMOCs that explain how outcomes were generated (see Appendix).^13^

CMOCs were examined both individually and in relation to one another, allowing us to construct an overarching explanatory account of how the model operates through layers of interdependent processes. Negative cases and boundary conditions were actively sought to identify where mechanisms were constrained or failed to produce intended outcomes. Findings were refined through team discussion and feedback from stakeholders, ensuring that the resulting explanations were both empirically grounded and practically meaningful.

### Reporting

The study was conducted and reported in accordance with realist evaluation principles and RAMESES reporting standards.^11^

### Ethics statement

Ethics approval ID 125999 was granted by the institutional review board of Western University, London, Ontario, Canada.

## RESULTS

### Overview of the realist explanatory account

Across stakeholder accounts, S.O.S emerged as a relational and adaptive infrastructure that makes care possible in contexts where conventional systems are experienced as unsafe, fragmented, or unresponsive. Understanding how this works requires looking at what services were delivered, at the conditions under which people were willing and able to engage with them, and how those conditions were sustained over time.

A linked set of realist explanations accounts for this. First, repeated, non-judgmental encounters in familiar settings build relational trust, enabling individuals who would otherwise avoid services to engage. Second, delivering care in locations that are accessible and acceptable to the clients reduces the practical and cognitive burden of participation, making engagement feasible. Third, the availability of a reliable and acceptable service in moments of crisis enables timely help-seeking, allowing harm reduction interventions to prevent serious outcomes and extend survival. Fourth, collaboration across sectors – grounded in trust and flexibility – allows providers to tailor responses and navigate system constraints, translating engagement into access to appropriate and coordinated care. Finally, an action-oriented approach that produces visible, practical results enables others to recognize the model’s value, generating legitimacy, mobilizing resources, and sustaining participation over time.

These realist explanations are not independent; they are intertwined and feedback on each other. The outcome of one intervention creates the context for another mechanism to act. Engagement depends on trust, feasibility depends on proximity, survival depends on timely access, and continuity depends on coordination. The model’s durability, in turn, depends on whether these practices generate visible value that others are willing to support.

Each realist explanation also reveals the conditions under which its mechanisms break down: where trust exists but no viable options are available; where proximity cannot overcome structural poverty or housing options; where harm reduction extends survival without access to longer-term care; where coordination meets institutional rigidity; and where sustained action relies on provider overstretching. These boundary conditions are part of the account because they show where relational and adaptive capacity meets structural limits that no outreach model can resolve on its own.

Together, the realist explanations and their limits suggest that S.O.S.’s effectiveness depends less on the availability of any particular service than on its capacity to reduce friction, enable timely response, and sustain relationships long enough for meaningful change to occur, while also revealing the structural conditions required for that change to endure.

### Realist explanation 1: Trust converts system avoidance into engagement with care

People experiencing homelessness, substance use, and prior negative encounters with formal services do not arrive at care as neutral actors. They arrive expecting stigma, dismissal, and bureaucratic gatekeeping; that is what they have learned to expect. SOS encounters them in a different context: familiar settings, informal interactions, no eligibility requirements, no compliance conditions. This context fosters trust and relationships that are the precursor for all that subsequently happens.

Providers show up repeatedly, in the same places. They remember names. They ask how someone is doing and mean it. One client described the moment recognition itself became evidence of care: *“From day one, we met, we talked, and the next time I seen her… she was like, hey Ed* [pseudonym], *how are you? … She knew who I was”* (P29). Being known disrupted an expectation of anonymity and dismissal that formal systems had spent years confirming. That disruption is a relational experience that shifts what feels safe enough to risk, not a program feature or a protocol.

Providers understood this sequencing clearly. As one described, *“We can’t do the medical work with them if we don’t have the connection… we’re… building personal rapport first”* (P31). Trust is something the model actively produces, encounter by encounter. And once produced, it travels. Trust carries a sense of safety into new service settings through what outreach providers called *“warm connections”* (P16), relational bridging that reduces the emotional risk of entering unfamiliar systems alongside someone who is already trusted.

The immediate outcome is engagement: genuine disclosure of need and willingness to accept support. Clients who had avoided services for years began to name what they needed. Referrals that would otherwise have been handed over on paper and forgotten were followed through. Short-term counseling functioned as a bridge: *“I provide ongoing short-term counseling… that is kind of the bridge to the next service… so we can kind of do the soft handoff”* (P22). Trust is the behavioural mechanism through which any subsequent care becomes possible at all.

### Realist Explanation 2: Proximity reduces friction and enables engagement with services

Accessing care requires more than knowing it exists. Conventional service systems are largely designed as if the gap between a referral and an attended appointment does not exist. As one SOS leader put it plainly: *“you hand someone a piece of paper… and you know it’s not coming back”* (P32). What happens when the system stops waiting and comes instead?

Closing this gap requires reducing the practical, cognitive, and emotional burden that engagement with care normally demands. In S.O.S, services are delivered in streets, encampments, shelters, and motels because that is where people already are. The resource mechanism is a reduction in the cumulative practical, cognitive, and emotional burden that engagement with care normally demands. One client described the specific texture of that burden: *“When you’re sleeping rough… without a cell phone… you have no idea what day of the week it is… that’s a big barrier… and they show up at the convenient places where we are”* (P30). When the effort required to initiate contact drops low enough, participation becomes possible for people for whom it was not before.

Proximity also transforms what a referral means in practice. SOS providers accompany clients, providing real-time reminders, helping complete paperwork, and walking alongside people through the early steps of engagement with new services. This is akin to a family member or friend who is physically present to absorb logistical complexity that would otherwise fall entirely on the person least equipped to carry it.

The immediate outcome is that people show up for appointments. Referrals convert into attended appointments. Paperwork gets completed. Transitions that stall elsewhere become navigable. And repeated contact under these conditions does more than produce single episodes of engagement; it builds the kind of familiarity and predictability that reinforces trust over time. Proximity makes participation feel feasible for people for whom the system has long felt otherwise.

### Realist Explanation 3: Accessible support enables timely help-seeking during periods of acute risk

The drug supply is unpredictable and increasingly toxic.^14-15^ People experiencing homelessness and active substance use face overdose risk, withdrawal, infections, and exposure to contaminated supplies in environments where formal services are often inaccessible, delayed, or experienced as too unsafe to approach. In those moments — the ones that are actually life-threatening — the question is not whether a service exists somewhere. It is whether a person will reach for it in time.

In these circumstances, timely help-seeking depends on whether support is experienced as safe, acceptable, and worth approaching during moments of acute vulnerability. Within SOS providers are known, non-judgmental, and genuinely approachable, clients are more likely to seek help in moments of acute risk rather than waiting, hiding, or managing alone. One provider described this as *“meeting people where they’re at”* (P22); a practical condition that reduces the hesitation and fear that ordinarily delay help-seeking in critical moments. The behavioural mechanism is the willingness to reach out, activated by a relationship with a service that has already proven itself safe to approach.

That willingness is what makes the interventions work. Naloxone distribution, drug checking, wound care, safer use supplies. These tools only prevent harm if someone reaches for them when danger is present. One client described exactly how that played out when using the drug testing equipment: *“You told me what was in the drugs… it had a fentanyl… really strong… if I hadn’t done that first… I would have been dead”* (P30). Fentanyl contamination is widespread.^16-18^ What was different was that this person sought out that information, in the moment it mattered, from a source they trusted enough to approach.

Harm reduction works through creating the conditions under which people are willing to access it when it counts.

The meta-outcome is survival: the avoidance of overdose deaths and severe acute complications. But the immediate outcome matters just as much. Stabilization, even when temporary, creates a window – people who survive acute crises remain reachable, and the opportunity for continued contact with services stays open. In a model where sustained relationship is the foundation of everything else, keeping that window open is what makes the rest of the work possible.

### Realist Explanation 4: Adaptive coordination enables access to appropriate care

Formal systems are not designed for the people SOS serves. Eligibility criteria, organizational mandates, professional scopes of practice, and institutional timelines are built around assumptions – of stable addresses, consistent communication, predictable needs – that do not hold for people experiencing homelessness and acute vulnerability. Navigating those systems on behalf of such clients requires the willingness of multiple actors, across sectors, to bend their usual processes in the same direction at the same time. What makes that willingness possible?

Through sustained contact and shared purpose, SOS has built a network of providers across health, social services, paramedicine, and community organizations who trust each other enough to act flexibly. That bridging of services is the behavioural mechanism. Internally, it manifests as role clarity and mutual confidence: *“We all trust each other to do what our skill set is and to know when it’s time to pass it off”* (P11). Decisions are made collectively, with different forms of expertise shaping responses regardless of professional status: *“We make these decisions together… whether you’re an unregulated provider at a charity or a regulated professional… we make these decisions together”* (P9). It is a flat structure by relationship, sustained because people have learned they can rely on one another.

Externally, that trust extends across institutional boundaries in ways that formal agreements rarely achieve. Providers described routinely *“working around the boundaries of their own organization… to make things work”* (P7), stretching their mandates in the direction of a shared client goal and not waiting for formal agreements to be drawn up and signed by all parties. While these workarounds appeared fragile, in practice, they enabled rapid action by relying on relationships rather than bureaucratic processes, allowing coordination to move at the speed the situation required. Stakeholders were also careful to note that this collaboration has not been *“all sunshine and roses”* (P1), requiring ongoing work to address tensions between organizations and navigate evolving team dynamics.

The immediate outcome is that clients gain timely access to services, exceptions, and coordinated responses that rigid systems would otherwise withhold. Care plans align across agencies rather than fragmenting at every handoff. Transitions that would stall in a siloed system become navigable when providers are in regular contact and willing to carry their part of the work. Cross-sector collaboration makes appropriate care accessible to people the system was not built to serve.

### Realist Explanation 5: Visible results build legitimacy and sustain support

Community responses to complex social problems often emerge in moments of urgency. Sustaining those responses over time, however, requires more than goodwill. People, organizations, and funders must become convinced that collective effort is producing meaningful results.

In Grey County, SOS emerged in response to a 24h notice of a crisis, looked at the systems that were supposed to address it, decided not to wait, and plan for a coordinated response. Existing structures were too slow, too fragmented, too constrained by their own mandates to move at the speed the situation demanded. What emerged was something more improvised and more durable: providers and community partners willing to take risks, cross boundaries, and act on what was needed right now rather than what was formally sanctioned. How did that improvised response become something that lasted?

Part of the answer is the behavioural mechanism of collective problem-solving under uncertainty. When people with different expertise, different organizational homes, and different formal roles come together around a shared and immediate problem, they drew on distributed knowledge that no single actor possesses. One team member captured it clearly: “*You get the people with the grit together and they’ll just do the work”* (P9). Role flexibility, real-time adaptation, and willingness to troubleshoot across organizational lines are what convert goodwill into actual solutions; the activation of collective capacity that flexible, trust-based collaboration makes available.

The resource mechanism is visibility. Action produces results, and results that can be seen change minds. As those early responses demonstrated what was possible, partners who had been skeptical became convinced. As one reflected, people *“came together and were like, wow, this really works. Let’s keep this going”* (P7). Seeing the model produce tangible outcomes in practice generates credibility that no proposal or planning document could have established in advance. That is how SOS makes the case for itself.

Together, collective problem-solving and the visibility of its results transform a crisis response into something institutionally legible and sustainable. The immediate outcome is that partners continue to show up because they have seen that showing up produces something real. Funders and institutions extend support because the model has demonstrated, in practice, what it can do. Cross-sector collaboration becomes normalized through demonstration. And the meta-outcome is that SOS becomes recognized as evidence that a different way of working is possible, which may be its most durable contribution of all.

### Boundary conditions and constraints: when the model hits system limits

While SOS demonstrates strong effectiveness across relational outreach, proximity-based care, harm reduction, coordination, and action-oriented practice, our analysis also identified consistent boundary conditions where these realist explanations reach their limits. Examining these boundary conditions helps clarify not only how SOS works, but under what circumstances its work is unable to translate relational and logistical gains into durable change. This section outlines these boundary conditions across explanations.

First, trust and relational engagement, although foundational, cannot by themselves produce forward movement when no viable system options exist. Participants described cases in which individuals remained connected to outreach providers and continued to accept contact, yet no housing placement, stabilization bed, or protective environment was available. In such instances, relational continuity functioned as accompaniment rather than transition. Similarly, where individuals refused care in the context of cognitive impairment or severe substance dependence, the voluntary nature of services limited what could be achieved. Trust enabled presence, but not protection.

Second, the realist explanation of proximity reducing friction has limits when structural poverty constrains exit pathways. Mobile services, escorted referrals, and active navigation increased uptake and follow-through, but participants emphasized that income inadequacy and housing market conditions repeatedly returned clients to homelessness. Even when individuals completed withdrawal management or temporary motel stays, discharge into an environment without affordable housing reversed gains. In this sense, proximity reduces access barriers but cannot resolve income–housing mismatches that lie outside the program’s control.

Third, timely help-seeking, while clearly life-preserving in many instances, also revealed a threshold beyond which voluntary, community-based care cannot contain risk. Participants described individuals whose acuity exceeded what outreach, medication support, or monitoring could stabilize. In the absence of appropriate longer-term or involuntary stabilization options, the system defaulted to crisis cycling. These cases illustrate that harm reduction extends survival windows but does not substitute for secure, longer-duration care infrastructures when required.

Fourth, adaptive coordination depends on reciprocity from receiving institutions. SOS successfully negotiated exceptions, accelerated referrals, and leveraged cross-sector relationships; however, gains often collapsed when clients entered rigid downstream systems governed by standardized timelines and eligibility rules. Hospital discharges to homelessness and short-term program limits repeatedly undermined stabilization.

Coordination proved most effective where institutions demonstrated flexibility; it stalled where systems reverted to transactional rather than relational logics.

Fifth, the model’s action orientation, which generated community legitimacy and sustained support, also produced workforce strain. The same visible responsiveness that built trust and political goodwill relied on staff and partner organizations extending themselves beyond formal mandates and funding envelopes. Participants described emotional exhaustion, demoralization from public stigma toward harm reduction, and reliance on under-resourced charitable partners. Thus, action functioned both as a strength and as a source of organizational fragility, where sustained performance depended on continued human overextension.

Across realist explanations, several cross-cutting constraints emerged. Housing shortages and income inadequacy limited transitions from stabilization to recovery. Institutional inflexibility functioned as a counter-mechanism, eroding relational gains made upstream. Public stigma toward help-seeking for harm reduction affected morale and threatened workforce sustainability. Finally, participants emphasized that the effectiveness of SOS is strongly tied to the relational capacity, risk tolerance, and adaptive judgment of specific individuals. This human dependency raises questions about sustainability and highlights that the model’s mechanisms are partly person-embedded.

Taken together, these boundary conditions suggest that SOS operates as a stabilizing and bridging system within a larger ecology that remains structurally misaligned. The initiative succeeds in extending safety, building connection, and navigating systems under constraint, but it cannot fully compensate for deficits in housing supply, income supports, long-term care capacity, or institutional flexibility. As a result, initiatives such as SOS are likely to remain necessary unless broader structural barriers and system constraints are addressed. Rather than signaling failure, these limits clarify where additional structural investment and policy adaptation would be required for relational and community-based models to achieve more durable outcomes.

## DISCUSSION

This study offers a realist explanation of how a community-based initiative functions as a relational and adaptive infrastructure in contexts where formal systems are experienced as fragmented, inaccessible, or unsafe. Rather than attributing outcomes to the delivery of discrete services, the analysis shows how engagement, stabilization, and continuity are produced through a sequence of five realist explanations. An action-oriented approach sustains these processes by generating visible results that build legitimacy and mobilize ongoing support. Together, these realist explanations operate as a linked configuration in which each creates the conditions for the next. Effectiveness, therefore, depends less on what services are available than on whether people are able and willing to engage with them, and whether providers can adapt them in response to need.

These findings extend prior realist and community health literature by specifying how commonly cited features such as trust, accessibility, and coordination function as generative mechanisms under conditions of instability.^19-21^ While previous work has emphasized the importance of outreach, harm reduction, and systems navigation, our analysis shows how proximity reduces the cumulative burden of engagement, making participation feasible in everyday conditions,^22-24^ and how the availability of acceptable, non-judgmental services enables timely help-seeking that directly affects survival.^25-26^ In addition, coordination is not simply a structural arrangement but depends on providers’ willingness to tailor and adapt services across organizational boundaries.^27-28^ By articulating how these realist explanations interact, the study contributes a more integrated account of how community-based models operate in practice.

The boundary conditions identified in this study are part of the explanation itself. They demonstrate that trust, proximity, and coordination can remain active yet be insufficient to produce intended outcomes when structural conditions are misaligned.^29-30^ For example, trust may enable sustained engagement without enabling forward movement in the absence of viable housing options; proximity may reduce barriers to access without altering trajectories shaped by income inadequacy; and harm reduction may extend survival without access to longer-term stabilization. Similarly, coordination depends on reciprocity from receiving institutions, and action-oriented practice depends on sustained human effort, both of which are vulnerable to system rigidity and workforce strain. These findings shift attention from whether interventions work to when and under what conditions their mechanisms can translate into durable change.

The program theory developed here suggests that similar realist explanations may operate in other settings where services are experienced as inaccessible or unsafe, particularly where repeated, non-judgmental contact, place-based delivery, and cross-sector relationships are feasible. However, transferability is constrained where structural supports – especially housing, income, and institutional flexibility – are lacking. The effectiveness of such models is therefore contingent not only on local relational capacity but also on broader system alignment. In this sense, community-based initiatives such as SOS function as stabilizing and bridging systems within a wider ecology,^31^ extending safety and enabling engagement while also revealing the limits of what can be achieved without structural change.

As with all realist analyses, the realist explanations presented represent theoretically informed interpretations rather than definitive causal claims. Mechanisms such as trust, willingness to seek help, and collaborative adaptation are not directly observable and may operate differently across contexts. While the program theory offers transferable propositions, further testing in other settings is needed to examine how the realist explanations presented here function under varying structural conditions.

### AI disclosure

ChatGPT was used during the preparation of this manuscript to support editing and refinement of language. Specifically, the authors used it to workshop abstract and call-out-box drafts, explore phrasing alternatives, and tighten prose for concision and flow. All conceptual development, argumentation, and final editorial decisions were made by the authors.

## Data Availability

All data produced in the present study are available upon reasonable request to the authors

